# Association of prior tuberculosis with altered cardiometabolic profiles of people with HIV: A comparative cross-sectional study in Uganda

**DOI:** 10.1101/2025.03.19.25324234

**Authors:** Joseph Baruch Baluku, Diana Karungi, Brenda Namanda, Sharon Namiiro, Shamim Katusabe, Angut Mary Madalen, Martin Nabwana, Ronald Olum, Felix Bongomin, Edwin Nuwagira, Grace Kansiime, Christian Kraef, Megan Shaughnessy, Joshua Rhein, David Meya

## Abstract

**Background:** Cardiovascular disease (CVD) is the leading cause of mortality among people with HIV (PWH), but the influence of co-infections like tuberculosis (TB) on CVD risk remains underexplored. We aimed to compare cardiometabolic profiles of PWH with and without prior TB to determine if prior TB is associated with distinct cardiometabolic profiles.

**Methods:** We conducted a comparative, cross-sectional study at a tertiary hospital in Kampala, Uganda. Participants were randomly sampled PWH aged ≥18 years on antiretroviral therapy. Specifically, we enrolled PWH with and without prior active TB (ratio of 1:1). Anthropometric measurements, blood pressure, fasting blood glucose (FBG), lipid profile, and glycated hemoglobin were assessed.

**Results:** A total of 396 participants were enrolled (196 TB survivors and 200 controls). TB survivors had higher median FBG (5.5 vs. 5.1 mmol/l, p<0.001) and a higher prevalence of DM (17.9% vs. 9.5%, p=0.015). However, they had lower body mass index (23.0 vs. 25.1 kg/m², p<0.001) and waist circumference (81.0 vs. 84.0 cm, p=0.026). TB survivors had higher HDL-c levels (1.0 vs. 0.8 mmol/l, p<0.001), lower LDL-c levels (2.7 vs. 3.1 mmol/l, p<0.001) and lower prevalence of dyslipidemia (81.7% vs. 96.5%, p<0.001). Prior TB was independently associated with higher prevalence of elevated FBG (adjusted prevalence ratio (aPR) 1.79, 95% CI 1.10-2.92) and DM (aPR 2.34, 95% CI 1.11-4.94), but decreased risk of obesity (aPR 0.42, 95% CI 0.20-0.88).

**Conclusion:** TB survivors with HIV exhibit a higher risk of DM but lower risk of obesity compared to those without a history of TB, indicating a need for blood glucose monitoring among TB survivors.

## BACKGROUND

Cardiovascular disease (CVD) is an emerging cause of significant morbidity and mortality among people with HIV (PWH) [1]. Antiretroviral therapy (ART) has increased the life-expectancy of PWH by suppressing HIV viral replication and reconstituting immune function [2]. As such, there has been a shift in the causes of death and morbidity among PWH from opportunistic infections to non-communicable diseases [3]. PWH are at a higher risk for myocardial infarction, coronary artery disease, stroke, pulmonary embolism, and peripheral artery disease compared to people without HIV infection [4–8]. Traditional CVD risk factors such as hypertension, obesity, dyslipidemia, and diabetes mellitus (DM) are thought to synergize with HIV-associated risk factors, including systemic inflammation and effects of ART, to heighten CVD risk among PWH [9–11]. However, the contribution of opportunistic infections to CVD risk is gaining prominence, particularly in low-income settings [9,12,13]. Low CD4 counts have been historically associated with cardiovascular events in PWH [11,12,14]. This suggests that PWH who are at risk of opportunistic infections may also be at higher risk of CVD, although it is unclear whether opportunistic infections directly modify traditional CVD risk factors.

Over 30% of PWH in sub-Saharan Africa are reported to be co-infected with TB [15]. Emerging evidence suggests that *Mycobacterium tuberculosis* (*Mtb*) is a risk factor for CVD across the entire spectrum of *Mtb* infection, albeit in predominantly HIV negative cohorts [16]. TB survivors are at a 51% higher risk of major adverse cardiovascular events; including non-fatal stroke, unstable angina and acute myocardial infarction and CVD is the leading cause of death in TB survivors [16,17]. The mechanisms by which *Mtb* contributes to an increased risk of CVD remain poorly understood. Our recent systematic review and meta-analysis revealed that individuals with active TB in Africa exhibit a high prevalence of traditional CVD risk factors [18]. Furthermore, our cross-sectional study in Uganda demonstrated that over 90% of TB patients— 56% of whom were PWH—have at least one CVD risk factor [19]. Whether prior TB specifically increases the prevalence of CVD risk factors in PWH remains uncertain. An increased risk would warrant heightened clinical surveillance for CVDs and their risk factors—including monitoring of blood lipids, blood pressure, blood glucose, and anthropometrics—among PWH who have completed TB therapy. We aimed to compare cardiometabolic profiles of PWH with and without prior TB to determine if prior TB is associated with cardiometabolic profiles.

## MATERIALS AND METHODS

### Study design and setting

This was a comparative, cross-sectional study to compare cardiometabolic profiles of PWH with and without prior TB. The study was conducted at Kiruddu National Referral Hospital (KNRH). KNRH is an urban tertiary teaching hospital located in Kampala, the capital city of Uganda. The HIV clinic at KNRH has more than 25,000 PWH under active follow-up.

### Study Population and sampling

Eligible participants were adult PWH (aged ≥18 years) receiving ART at the HIV clinic at KNRH. Potential participants were identified from the HIV care database at KNRH and disaggregated by TB treatment history. Participants in the prior TB group were PWH who had any lifetime history of treatment for active bacteriologically confirmed TB by the GeneXpert assay or lateral flow urine lipoarabinomannan test (Alere Determine™ TB LAM Ag) as defined by the World Health Organization [20]. We excluded PWH who were on current TB treatment. Thereafter, potential participants were randomly selected from each of the two groups using a random computer-generated sequence. Selected potential participants were invited to participate in the study after a brief screen for eligibility via a phone call. Those willing to participate and eligible were invited to KNRH to provide written consent and undergo study measurements after an overnight fasting period.

### Study measurements

Trained research assistants administered a questionnaire to eligible participants for socio-demographic characteristics, medical history, any history of cigarette smoking and alcohol use. Alcohol use was graded using the CAGE questionnaire. The CAGE questionnaire is a set of 4 simple questions: “Have you ever: (1) felt the need to Cut down your drinking; (2) felt Annoyed by criticism of your drinking; (3) had Guilty feelings about drinking; and (4) taken a morning Eye opener [21]. HIV and TB treatment history were extracted from the HIV treatment database and TB unit register at KNRH. Participants underwent anthropometric measurements of their weight, height, mid upper arm, neck, waist and hip circumference using a weighing scale (Seca 760®), stadiometer (Seca 213®), and tape measure respectively. The body mass index (BMI) was calculated using the formula; BMI = weight (kilograms)/height (metres)^2^. Using a battery powered digital blood pressure (BP) machine (Omron®, Hem 7120), the office BP was taken on two separate occasions, 20 minutes apart. The average BP of the two measurements was considered as the participant’s BP. A study nurse drew 4 milliliters (mls) of blood, which was tested for the fasting blood glucose (FBG), glycated haemoglobin (HbA1c), serum uric acid, and fasting lipid profile. The FBG was measured using a point-of-care glucometer (Accu-Chek®).

The blood lipids (triglycerides, total cholesterol, low density lipoprotein cholesterol (LDL-c), and high-density lipoprotein cholesterol (HDL-c)) were estimated using the Cobas® 6000 analyzer series (Roche Diagnostics, USA).

### Study Outcomes

The primary outcome measure was the frequency of each of hypertension, dyslipidemia, elevated FBG, DM, central obesity, obesity, and high BMI among PWH with and without prior TB. Secondary outcomes were elevation in each of systolic BP, diastolic BP, HbA1c, total cholesterol, LDL-c, HDL-c, triglycerides, and waist circumference. Hypertension was defined as elevated systolic BP ≥ 140 mmHg and/or diastolic ≥ 90 mmHg and/or prescribed use of anti-hypertension medication [22]. Dyslipidemia was defined as an elevated total cholesterol of > 5.0 mmol/l and/or LDL-c of > 4.14 mmol/l, and/or triglyceride level of ≥1.7 mmol/l, and/or low HDL-c of <1.03 mmol/l for men and <1.29 mmol/l for women [23]. The FBG was considered elevated if ≥5.6 mmol/l while an HbA1c ≥ 5.7% was considered elevated. DM was defined as FBG ≥7.0 mmol/l and/or HbA1c ≥6.5% and/or use of anti-hyperglycemic agents [24]. Central obesity was defined as an elevated waist circumference of ≥ 102/88 cm and/or a waist-hip ratio of ≥0.90/0.85 in males/females, respectively [25]. High BMI was a BMI ≥ 25 kilograms/meters^2^ while obesity was a BMI of ≥ 30 kg/m^2^.

### Data analysis and sample size estimation

In estimating the sample size, we assumed the prevalence of CVD risk factors among PWH without prior TB as 23% for high BMI, 12% for central obesity, 16% for hypertension and 1% for DM based on prevalence data among PWH in Uganda [26]. For PWH with prior TB we assumed a prevalence of 8% for high BMI, 39% for central obesity, 41% for hypertension, and 7% for DM [19]. Using the OpenEpi sample size calculator [27], the comparison for DM gave the largest sample size of approximately 400 (200 PWH TB survivors and 200 PWH without prior TB). The calculation assumed a study power of 80% and a two-sided significance level of 5%.

Categorical variables were compared between PWH with and without prior TB using Pearson’s chi-square test or Fisher’s exact test as appropriate. Continuous variables were compared between PWH with and without prior active TB infection using independent t-test or Mann-Whitney U test as appropriate. The frequency of hypertension, dyslipidemia, elevated FBG, DM, central obesity, obesity and high BMI were compared using Pearson’s chi-square test among PWH with and without prior active TB. We calculated and compared the 10-year CVD risk scores among PWH with and without prior TB using the Framingham’s risk score (FRS) [28].

Individuals were classified as having a low CVD risk if FRS <10%; moderate CV risk if FRS is 10-20% or high CV risk if FRS ≥20%.

To determine if prior TB is independently associated with the cardiometabolic profiles, we constructed multivariable Modified Poisson regression models with robust standard errors for factors associated with each of hypertension, dyslipidemia, elevated FBG, DM, central obesity, obesity, and high BMI – a total of seven models. Similar models were constructed for each of elevation in systolic BP, diastolic BP, HbA1c, total cholesterol, LDL-c, HCL-c, triglycerides, and waist circumference. In constructing these models, variables with p-values less than 0.05 at bivariate analysis were included in the final multivariable models. We then intentionally controlled for prior TB in each model to determine if prior TB is independently associated with each of the above outcome variables. Statistical significance was set at p<0.05 for all analyses. All analyses were performed using STATA (version 18.0).

### Ethics committee approval and consent to participate

All study procedures were conducted in accordance with the Declaration of Helsinki. The study was approved by the Mildmay Uganda Research Ethics Committee (MUREC-2023-240), and the Uganda National Council of Science and Technology (HS2991ES) prior to participant recruitment. Study participants provided written informed consent before study procedures were performed.

## RESULTS

We screened 599 records of PWH and enrolled 396 PWH (196 TB survivors and 200 without prior TB). Among those who were not enrolled, 183 had disconnected phone numbers and could not be contacted, 13 did not consent to participate, 3 were currently on TB re-treatment and 4 PWH TB survivors were found to have had clinically diagnosed TB (no bacteriological confirmation). **Figure 1** shows the study flow diagram.

**Figure 1:**
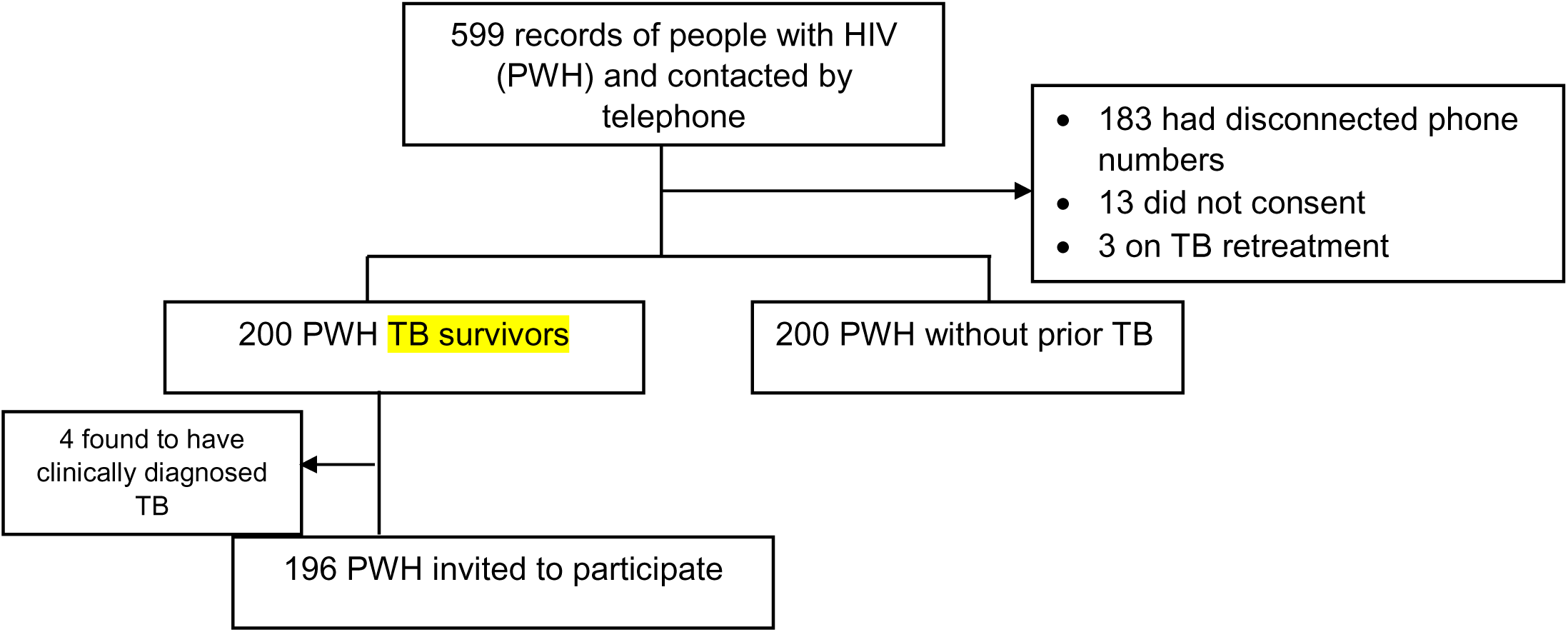
Study flow diagram

### Characteristics of the study population

The mean age (standard deviation, SD) of the entire study population was 41.5 (12.2) years (**table 1)**. Among PWH TB survivors, 22 (11.3%) had been treated more than once, 161 (82.1%) had TB confirmed by GeneXpert and 35 (17.9%) by urine lipoarabinomannan. Further, 12 (6.1%) had had drug resistant TB, 190 (96.9%) had had documented TB cure, and the median (IQR) duration since TB cure/treatment completion was 11.8 (5.2-28.6) months. Table 1 compares characteristics of both study groups.

**Table 1:** Characteristics of people with HIV with and without prior tuberculosis.

| Characteristic | Total<br>N=396 | People with HIV<br>without TB history<br>N=200 | People with HIV with<br>TB history<br>N=196 | p-value |
| --- | --- | --- | --- | --- |
| **Age (Years), Mean (SD)** | 41.5 (12.2) | 41.3 (12.9) | 41.7 (11.5) | 0.720 |
| **Male Sex** | 185 (46.7) | 74 (37.0) | 111 (56.6) | <0.001 |
| **Tribe** |  |  |  | 0.170 |
| Baganda | 253 (64.2) | 119 (59.8) | 134 (68.7) |  |
| Banyakitara | 81 (20.6) | 47 (23.6) | 34 (17.4) |  |
| Others | 60 (15.2) | 33 (16.6) | 27 (13.8) |  |
| **Urban Residence** | 279 (70.5) | 148 (74.0) | 131 (66.8) | 0.120 |
| **Duration at current residence (Months), Median (IQR)** | 7.0 (3.0-20.0) | 6.0 (3.0-15.0) | 10.0 (4.0-22.5) | 0.008 |
| **Married** | 176 (44.4) | 87 (43.5) | 89 (45.4) | 0.700 |
| **At least Secondary school education level** | 192 (48.5) | 90 (45.0) | 102 (52.0) | 0.160 |
| **Unemployed** | 67 (16.9) | 28 (14.0) | 39 (19.9) | 0.120 |
| **Currently on Trimethoprim/Sulfamethoxazole prophylaxis** | 100 (25.3) | 32 (16.0) | 68 (34.9) | **<0.001** |
| **Drugs in antiviral therapy (ART) Regimen** |  |  |  |  |
| Tenofovir | 380 (96.0) | 188 (94.0) | 192 (98.0) | 0.071 |
| Lamivudine | 392 (99.0) | 196 (98.0) | 196 (100.0) | 0.120 |
| Abacavir | 6 (1.5) | 5 (2.5) | 1 (0.5) | 0.220 |
| Zidovudine | 6 (1.5) | 4 (2.0) | 2 (1.0) | 0.690 |
| Efavirenz | 21 (5.3) | 13 (6.5) | 8 (4.1) | 0.370 |
| Dolutegravir | 365 (92.2) | 182 (91.0) | 183 (93.4) | 0.460 |
| Protease inhibitor (Lopinavir, Ritonavir, or Atazanavir) | 13 (4.4) | 4 (2.0) | 9 (4.5) | 0.259 |
| **Duration on ART (Years), Median (IQR)** | 4.0 (1.7-10.0) | 6.0 (3.0-11.5) | 3.0 (1.0-8.0) | **<0.001** |
| **History of ART default** | 40 (10.2) | 20 (10.1) | 20 (10.3) | 0.950 |
| **CD4 count at HIV diagnosis, (cells/UI), Median (IQR)** | 237.5 (97.5-471.0) | 289.0 (116.0-516.0) | 200.0 (74.0-406.0) | 0.051 |
| **Viral load suppression (threshold of <1000 copies/mL)** | 309 (94.8) | 176 (96.2) | 133 (93.0) | 0.200 |
| **At least one current opportunistic infection\*** | 23 (5.8) | 1 (0.5) | 22 (11.2) | **<0.001** |
| **Clinical stage of HIV at time of HIV diagnosis** |  |  |  | **<0.001** |
| Stage I | 176 (44.4) | 159 (79.5) | 17 (8.7) |  |
| Stage II | 38 (9.6) | 36 (18.0) | 2 (1.0) |  |
| Stage III/IV | 182 (46.0) | 5 (2.5) | 177 (90.3) |  |
| **At least one past opportunistic infection (except Lower Respiratory Tract Infection)** | 58 (14.6) | 27 (13.5) | 31 (15.8) | 0.510 |
| **Pre-existing cardiometabolic disease** |  |  |  |  |
| Diabetes | 25 (6.3) | 14 (7.0) | 11 (5.6) | 0.680 |
| Hypertension | 66 (16.7) | 38 (19.0) | 28 (14.3) | 0.230 |
| Dyslipidemia | 1 (0.3) | 1 (0.5) | 0 (0.0) | 1.000 |
| Renal disease | 1 (0.3) | 1 (0.5) | 0 (0.0) | 1.000 |
| **Alcohol use** |  |  |  | **0.570** |
| Never used alcohol | 158 (40.1) | 85 (42.5) | 73 (37.6) |  |
| Formerly used alcohol (>6 months) | 118 (29.9) | 56 (28.0) | 62 (32.0) |  |
| Currently uses alcohol (within past 6 months) | 118 (29.9) | 59 (29.5) | 59 (30.4) |  |
| **CAGE<sup>#</sup>** |  |  |  | **0.002** |
| 0-1 | 372 (94.4) | 196 (98.0) | 176 (90.7) |  |
| 2-4 | 22 (5.6) | 4 (2.0) | 18 (9.3) |  |
| **Smoking** |  |  |  | **0.210** |
| Never smoked | 314 (80.1) | 165 (82.9) | 149 (77.2) |  |
| Formerly smoked (>6 months) | 51 (13.0) | 20 (10.1) | 31 (16.1) |  |
| Currently smokes (within past 6 months) | 27 (6.9) | 14 (7.0) | 13 (6.7) |  |
| **Pack years of smoking, Median (IQR)** | 1.8 (0.8-3.2) | 1.0 (0.5-2.8) | 2.0 (0.8-6.0) | 0.210 |
| **Any family history of cardiovascular disease in first degree relative<sup>§</sup>** | 177 (44.7) | 79 (39.5) | 98 (50.0) | **0.036** |
| **Specific family history of cardiovascular disease in first degree relative** |  |  |  |  |
| Diabetes mellitus | 82 (46.3) | 30 (38.0) | 52 (53.1) | 0.050 |
| Hypertension | 132 (74.6) | 62 (78.5) | 70 (71.4) | 0.300 |
| Kidney disease | 7 (4.0) | 1 (1.3) | 6 (6.1) | 0.130 |
| Stroke | 13 (7.3) | 6 (7.6) | 7 (7.1) | 1.000 |
| Obesity | 16 (9.0) | 5 (6.3) | 11 (11.2) | 0.300 |
| Heart failure | 5 (2.8) | 1 (1.2) | 4 (4.1) | 0.380 |
| **Conjunctival Pallor** | 53 (13.4) | 23 (11.5) | 30 (15.3) | 0.270 |
| **Temperature (°C), Mean (SD)** | 35.9 (0.7) | 36.0 (0.6) | 35.9 (0.7) | 0.092 |
| **Pulse rate (beats per minute), Mean (SD)** | 76.4 (12.8) | 76.1 (12.4) | 76.6 (13.3) | 0.710 |
| **Abnormalities on chest auscultation** | 32 (8.1) | 6 (3.0) | 26 (13.3) | **<0.001** |
| **SPO2 (%), Mean (SD)** | 96.5 (2.7) | 96.9 (2.0) | 96.1 (3.3) | **0.004** |
| **Respiratory rate (breaths/min), Mean (SD)** | 18.5 (2.6) | 18.5 (2.6) | 18.6 (2.6) | 0.560 |

\*oral candidiasis, fungal nail infection, herpes zoster, recurrent upper respiratory tract infection (sinusitis, pharyngitis, tonsillitis, otitis media), and lower respiratory tract infections (especially pneumonia). <sup>#</sup> score for hazardous alcohol use (C- cut down, A – anger, G –guilt, E – eye opener), <sup>§</sup> includes any of Diabetes mellitus, Hypertension, Kidney disease, Stroke, Obesity and Heart failure. Bolded p-values indicate a statistically significant result

### A comparison of cardiometabolic profiles between PWH with and without prior TB

PWH with a history of TB had a significantly lower median body weight (60.0 kg vs 64.0 kg, p=0.011), BMI (23.0 kg/m^2^ vs. 25.1 kg/m^2^, p<0.001), and waist circumference (81.0 cm vs. 84.0 cm, p=0.026) than PWH without a history of TB. The prevalence of high BMI (25.6% vs. 51.0%, p<0.001) and obesity (7.7% vs. 24.5%, p<0.001) was also significantly lower in PWH with a history of TB.

However, PWH with a history of TB had higher median FBG levels (5.5 mmol/l vs. 5.1 mmol/l, p<0.001), higher prevalence of elevated FBG (≥5.6 mmol/l) (49.5% vs. 27.2%, p<0.001), and DM (17.9% vs. 9.5%, p=0.015) than PWH without a history of TB.

Additionally, PWH with a history of TB had higher mean HDL-c levels (1.1 mmol/l vs. 0.9 mmol/l, p<0.001) and lower mean LDL-c levels (2.8 mmol/l vs. 3.1 mmol/l, p=0.002) than PWH without a history of TB. The prevalence of dyslipidemia was also significantly lower in PWH with a history of TB (81.7% vs. 96.5%, p<0.001).

There were no significant differences in the measurements of central obesity, waist-hip ratio, blood pressure, hypertension, total cholesterol, uric acid or triglyceride levels between the two groups (**Table 2)**.

**Table 2:**
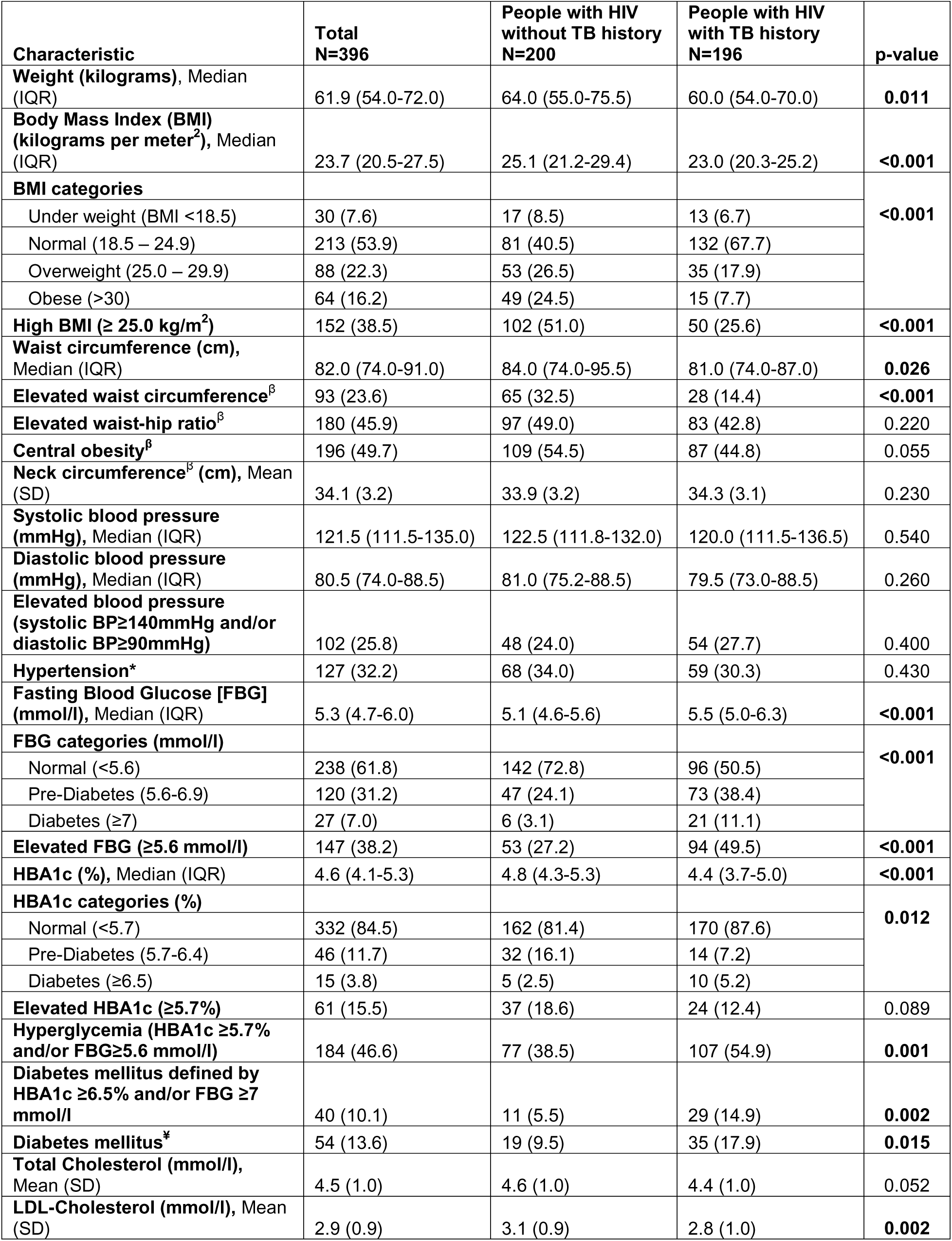
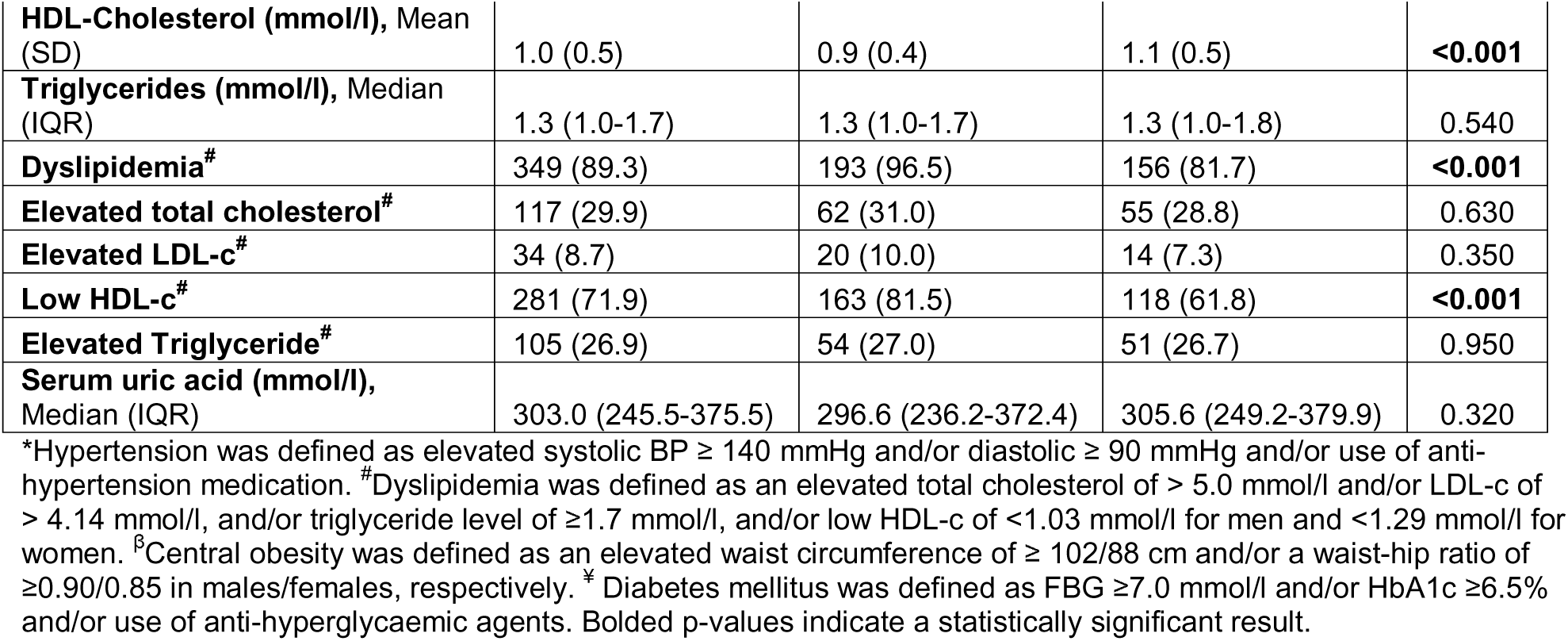
Cardiometabolic profiles of people with HIV with and without prior tuberculosis.

### Ten-year CVD risk among PWH with and without prior TB

The median ten-year CVD risk score was higher among PWH with TB history than those without (1.4% (IQR: 0.4 – 4.4) vs. 1.2% (0.3 – 3.0), p = 0.180); without reaching statistical significance. Similarly, a higher proportion of those with TB history had moderate or high CVD risk compared to those without TB history (19 (10.1%) vs. 13 (6.5%), p = 0.202). (**supplementary Table 9**)

### Association between prior TB and cardiometabolic profiles of PWH

In adjusted analyses, a history of TB was independently associated with a higher risk of elevated FBG (adjusted prevalence ratio (aPR) 1.79, 95% CI 1.10-2.92, p = 0.020) and DM (aPR 2.34, 95% CI 1.11-4.94, p = 0.025), but a decreased risk of obesity (adjusted risk ratio (aRR) 0.42, 95% CI 0.20-0.88, p = 0.021) **(table 3 and Supplementary tables 1 – 8).** Other factors associated with the individual cardiometabolic risk factors are described in the supplementary material.

**Table 3:**
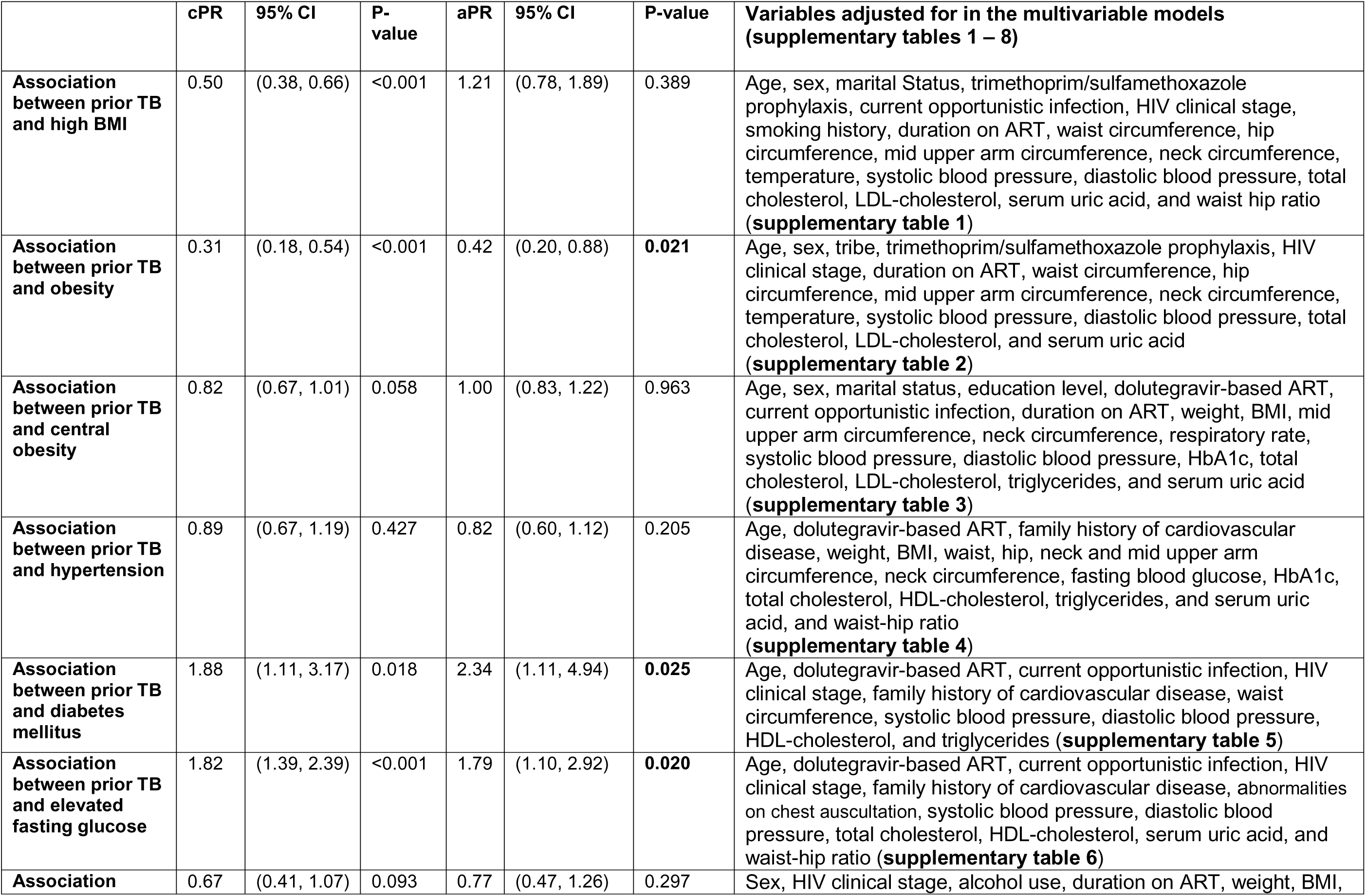
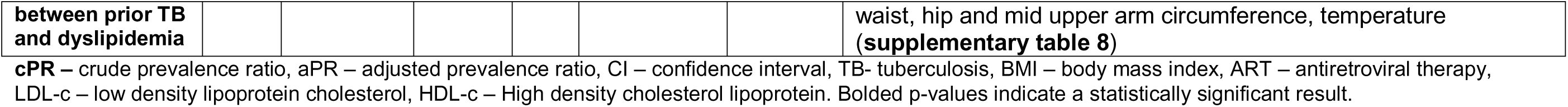
Association between prior TB and individual cardiometabolic profiles.

## DISCUSSION

We aimed to determine whether prior TB disease is independently associated with cardiometabolic profiles of PWH. PWH with a history of TB were found to have a significantly higher prevalence of DM and elevated FBG levels. This could be attributed to chronic inflammatory states induced by TB, which are known to disrupt insulin signaling and glucose metabolism, leading to insulin resistance—a precursor to DM [29]. These findings are consistent with other studies that have highlighted persistent hyperglycemia/DM among patients with TB although these studies are limited by a short follow up time [30]. The TB survivors in our study were enrolled at a median of almost a year after TB cure and rates of pre-existent known DM were similar between the two groups prior to study enrolment. Similar to our findings, Pearson and colleagues reported an elevated risk of DM among TB survivors followed for even a longer time (4 years) [31]. This suggests that the observed dysglycemic states are persistent and perhaps proceed the active TB period. The implication for clinical practice is that there is a need for increased screening, monitoring, and management of blood glucose levels among TB survivors to prevent the onset of DM and its complications. A critical question to address is the suitability of screening tests for detecting DM among TB survivors. Our study indicates that HbA1c, when compared to FBG, significantly underestimates the prevalence of both pre-diabetes and DM among TB survivors by factors of five (7.2% vs. 38.4%) and two (5.2% vs. 11.1%), respectively. This discrepancy was not observed among PWH who had never had TB. Although HbA1c is known to underestimate hyperglycemia in PWH generally [32], our findings suggest that a history of TB further exacerbates the discrepancy between HbA1c and FBG measurements. Therefore, HbA1c may not be a suitable test for DM screening in TB survivors.

Our findings also indicated a lower prevalence of obesity among TB survivors as well as an independent association between prior TB with low rate of obesity. A low prevalence of obesity might be linked to the reduced food intake due to altered appetite mediators and the subsequent weight loss associated with active TB infection [33]. While weight gain is observed in the early phases of TB treatment, there are minimal changes after TB cure probably due to lasting alterations in energy metabolism or changes in body composition that are not fully reversed after TB cure [34,35]. This aspect of TB’s legacy on body weight could potentially confer some protective effect against the traditional cardiovascular risk factors associated with obesity such as dyslipidemia. Our mediation analysis suggests that prior TB lowers LDL-c levels partly through its effect on BMI (supplementary figure 1). Taken together, it appears that TB survivors are at risk of presumed non-autoimmune DM despite normal BMI – a phenotype observed in Ugandan patients with DM [36].

Contrary to the increased risk of DM, our study found that TB survivors had a more favorable lipid profile, characterized by higher HDL-c levels and lower LDL-c levels. While these findings may suggest a lower risk of CVD, the clinical interpretation requires caution, as the overall impact of these altered lipid profiles in the context of HIV and TB co-morbidity is not fully understood. Although HDL-c plays a crucial role in transporting cholesterol from peripheral tissues back to the liver for excretion and possesses antioxidant, anti-thrombotic, and anti-inflammatory properties, elevated levels of HDL-c have paradoxically been associated with an increased risk of CVD [37]. Our analyses show an association between an increase in HDL-c with a higher prevalence of DM and hypertension (supplementary material). Therefore, elevation in HDL-c may not portend protection against CVD

This study has some limitations. The cross-sectional design limits our ability to establish temporality between prior TB and the observed cardiometabolic profiles. It is possible that survivorship bias is a factor here. PWH who have also had TB and survived might represent a unique group whose pre-existing cardiometabolic health contributed to their TB survival, rather than TB itself altering their metabolic profiles. However, DM and lower BMI are known to increase the risk of death from TB [38,39]. Therefore, it’s unlikely that the observed high prevalence of these conditions would have favored the survival of this population. Second, our study population was drawn from a single tertiary hospital in an urban setting, which may limit the generalizability of our findings to other regions or rural populations. Further, although we controlled for potential confounders in the multivariable models (see supplementary tables), we might have failed to control for other potential confounders such as genetic variations and dietary habits. Lastly, we did not perform the oral glucose tolerance test (OGTT), which would otherwise complement our assessment of glucose metabolism. However, while some studies suggest that the OGTT performs better than the HbA1c and FBS in PWH [32], other studies show that FBS [40] and HbA1c [41] perform better than the OGTT. Therefore, there is no clear consensus on the ideal test. Despite these limitations, our study has strengths. It is one of the few studies to comprehensively assess cardiometabolic profiles in PWH with a history of TB in a sub-Saharan African setting, contributing valuable data to an under-researched area. The use of well-established and standardized measurements for cardiometabolic outcomes, including extensive anthropometric assessment, HbA1c, FBG, lipid profiles, and blood pressure, enhances the reliability of our findings.

## CONCLUSION

This study demonstrates that PWH who have survived TB exhibit distinct cardiometabolic profiles compared to those without a history of TB. Specifically, TB survivors are at a significantly higher risk of developing DM and have elevated FBG levels, despite having a lower prevalence of obesity. The findings emphasize the need for enhanced monitoring and management of blood glucose levels in TB survivors with HIV, even in the absence of traditional risk factors like obesity.

## Data Availability

The datasets used and/or analysed during the current study are available from the corresponding author on reasonable request.

## Acknowledgements

None

## Authors’ contribution

JBB – conceptualizing, methodology, data accrual, data analysis, visualization, writing draft manuscript, revising manuscript, final approval.

MN - Methodology, data analysis, visualization, writing draft manuscript, revising manuscript, final approval.

DK, SN, SK, AMM - Methodology, data accrual, interpretation of results, revising manuscript, final approval.

RO, FB, EN, GK, CK, MS - Methodology, data visualization, revising manuscript, final approval.

JR, DM – Conceptualization, Methodology, data visualization, revising manuscript, final approval.

