## Supplementary material for "Association of prior tuberculosis with altered cardiometabolic profiles of people with HIV: A comparative cross-sectional study in Uganda"

**OTHER FACTORS ASSOCIATED WITH INDIVIDUAL CARDIOMETABOLIC PROFILES AMONG PWH**

**Factors associated with obesity and High BMI among people with HIV (PWH)**
Females (aPR = 2.40, 95% CI 1.07 – 5.38, p= 0.034) were more likely to have obesity while those with HIV clinical stage II (aPR= 0.45, 95% CI 0.23 – 0.86, p = 0.017) were less likely to have obesity compared to those with stage I. A unit increase in age (aPR = 1.03, 95% CI 1.01 – 1.05, p=0.011), hip circumference (aPR = 1.03, 95% CI 1.01 – 1.05, p = 0.010), and waist circumference (aPR = 1.03, 95% CI 1.02 – 1.04, p<0.001) were associated with obesity.

Further, Females (aPR = 1.54, 95% CI 1.11 – 2.15, p= 0.010), and married people (aPR = 1.30, 95% CI 1.03 – 1.64, p = 0.026) were more likely to have high BMI. PWH on Trimethoprim/Sulfamethoxazole prophylaxis (aPR = 0.56, 95% CI 0.38 – 0.92, p= 0.019) and those with HIV clinical stage II (aPR= 0.65, 95% CI 0.45 – 0.94, p = 0.021) and stage III/IV (aPR = 0.60, 95% CI 0.38 – 0.95, p = 0.030) were less likely to have high BMI. Additionally, a unit increase in the hip circumference (aPR = 1.03, 95% CI 1.10 – 1.05, p = 0.005), mid-upper arm circumference (aPR = 1.10, 95% CI 1.05 – 1.16, p<0.001), and serum uric acid levels (aPR = 1.00, 95% CI 1.00 – 1.00, p = 0. 048) were associated with high BMI **(supplementary table 1 and 2)**.

**Factors associated with central obesity among PWH**
Females (aPR = 1.78, 95% CI 1.28 – 2.49 p= 0.001), married people (aPR = 1.29, 95% CI 1.05 – 1.58, p= 0.015), and those with at least one previous opportunistic infection (aPR= 1.28, 95% CI 1.03 – 1.59, p=0.027) were more likely to have central obesity. A unit increase in age (aPR= 1.02, 95% CI 1.10 – 1.03, p=0.001), neck circumference (aPR = 1.04, 95% CI 1.00 – 1.08, p= 0.049), respiratory rate (aPR = 1.05, 95% CI 1.02 = 1.09, p = 0.004), HbA1c (aPR = 1.06, 95% CI 1.00 – 1.12, p= 0.036) and LDL-c (aPR = 1.12, 95% CI 1.01 – 1.25, p = 0.033) was associated with central obesity **(supplementary table 3).**

**Factors associated with hypertension and elevated BP among PWH**
An increase in age (aPR = 1.03, 95% CI 1.02 – 1.05, p<0.001), hip circumference (aPR = 1.02, 95% CI 1.00 – 1.04, p =0.017), HBA1c (aPR = 1.14, 95% CI 1.07 – 1.21, p<0.001), and HDL-c (aPR = 1.46, 95% CI 1.17 – 1.82, p=0.001) was associated with hypertension **(supplementary table 4)**.
Factors associated with elevated BP were age (aPR= 1.03, 95%CI 1.01 – 1.04, p<0.001), family history of CVD in a first degree relative (aPR = 1.62, 95% CI 1.13 – 2.33, p=0.009), an increase in the hip circumference (aPR = 1.03, 95% CI 1.01 – 1.05, p=0.002), HbA1c (aPR = 1.14, 95% CI 1.07 – 1.21, p<0.001) and HDL-c (aPR= 1.70, 95% CI 1.35 – 2.15, p<0.001). Conversely, an increase in the BMI was associated with a lower prevalence of elevated BP (aPR = 0.92, 95% CI 0.85 – 0.99, p = 0.025).

**Factors associated with diabetes mellitus, elevated FBG and HbA1c among PWH**

PWH with at least one current opportunistic infection (aPR= 2.70, 95% CI 1.35 – 5.39, p=0.005) were more likely to have DM and an increase in the HDL-c was associated with DM (aPR = 1.51, 95% CI 1.15 – 2.00, p=0.003). A dolutegravir-based regimen was associated with a lower prevalence of DM (aPR = 0.19, 95% CI 0.11 – 0.31, p<0.001).

Similarly, PWH with at least one current opportunistic infection (aPR= 1.57, 95% CI 1.12 – 2.19, p=0.009) were more likely to have elevated FBG. An increase in the HDL-c (aPR = 1.27, 95% CI 1.05 – 1.54, p=0.013) and serum uric acid (aPR = 1.00, 95% CI 1.00 – 1.00, p= 0.016) was also associated with elevated FBG.

An increase in the hip circumference (aPR = 1.01, 95% CI 1.00 – 1.03, p= 0.029), systolic BP (aPR= 1.01, 95% CI 1.00 – 1.02, p=0.012) and SPO2 (aPR = 1.11, 95% CI 1.01 – 1.23, p=0.038) were associated with an elevated HbA1c (**Supplementary tables 5 – 7)**.

**Factors associated with dyslipidemia among PWH**

Females had higher prevalence of dyslipidemia (aPR = 1.12, 95% CI 1.02 – 1.22 p= 0.020) while PWH reporting current alcohol use had lower prevalence of dyslipidemia (aPR = 0.90, 95% CI 0.82 – 0.98, p=0.023). An increase in weight (aPR = 1.01, 95% CI 1.00 – 1.02, p=0.003) and hip circumference (aPR = 1.00, 95% CI 1.00 – 1.00, p = 0.032) were associated with dyslipidemia (**supplementary table 8)**. Liner regression analyses showed that the relationship between previous TB and LDL-c levels is partially mediated by BMI: indirect effects (ab= -0.085, 95% CI -0.142 – -0.029, p = 0.003), and direct effects (c’ = -0.217, 95% CI -0.405 – -0.029, p = 0.023) (**Figure 1)**.

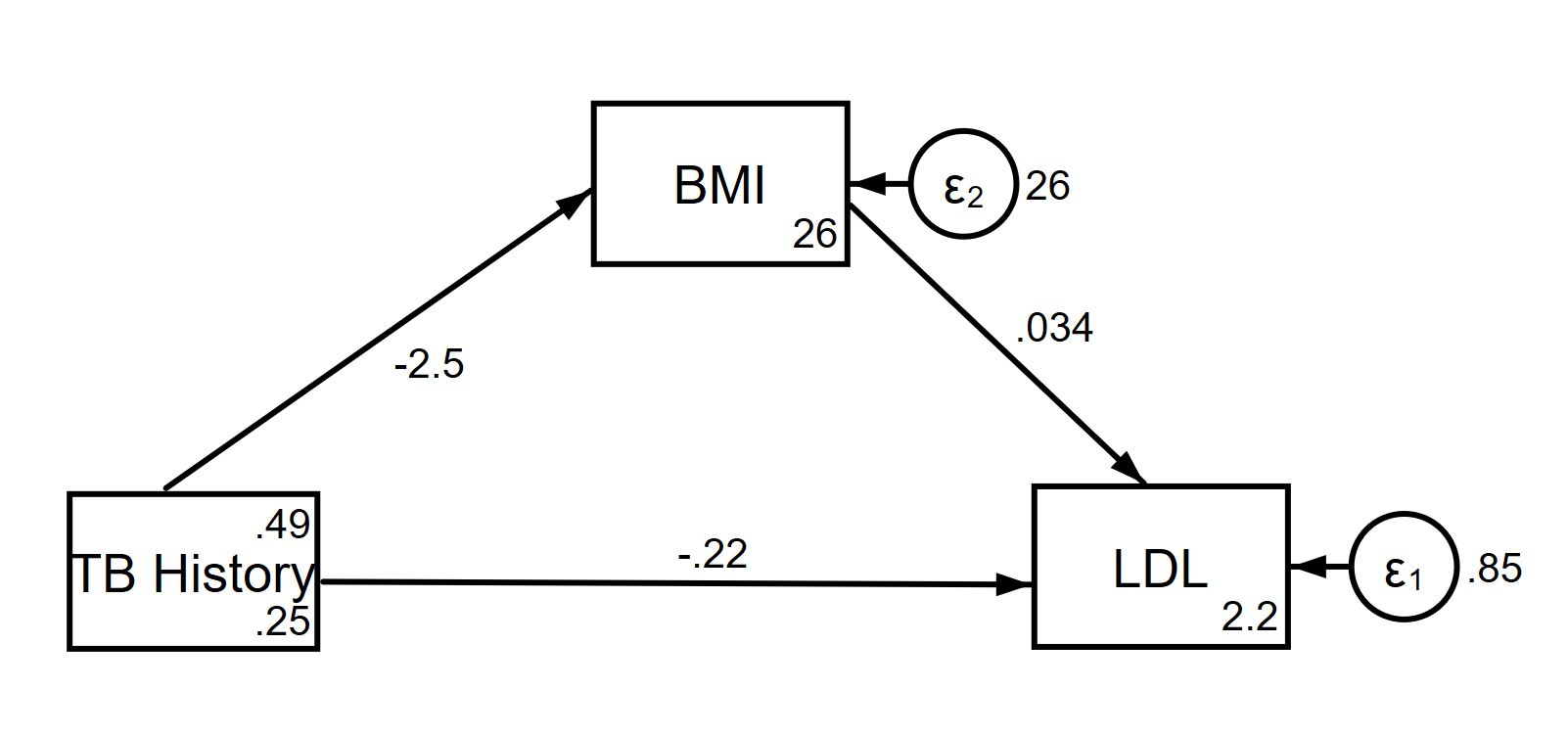

**Figure 1: Path diagram showing the mediation effect of body mass index on the relationship between previously treated active TB and low-density lipoprotein cholesterol**

**Table 1: Factors associated with high body mass index among people with HIV at an urban tertiary hospital in Uganda**

| **Characteristic** | **cPR** | **95% CI** | **P-value** | **aPR** | **95% CI** | **P-value** |
| --- | --- | --- | --- | --- | --- | --- |
| **TB history** |  |  |  |  |  |  |
| No | 1.00 |  |  | 1.00 |  |  |
| Yes | 0.50 | (0.38, 0.66) | <0.001 | 1.21 | (0.78, 1.89) | 0.389 |
| **Sex** |  |  |  |  |  |  |
| Male | 1.00 |  |  | 1.00 |  |  |
| Female | 1.85 | (1.40, 2.44) | <0.001 | 1.54 | (1.11, 2.15) | 0.010 |
| **Marital Status** |  |  |  |  |  |  |
| Not Married | 1.00 |  |  | 1.00 |  |  |
| Married | 1.42 | (1.11, 1.82) | 0.006 | 1.30 | (1.03, 1.64) | 0.026 |
| **Currently on Trimethoprim/Sulfamethoxazole prophylaxis** |  |  |  |  |  |  |
| No | 1.00 |  |  | 1.00 |  |  |
| Yes | 0.39 | (0.26, 0.61) | <0.001 | 0.59 | (0.38, 0.92) | 0.019 |
| **At least one current opportunistic infection** |  |  |  |  |  |  |
| No | 1.00 |  |  | 1.00 |  |  |
| Yes | 0.22 | (0.06, 0.82) | 0.024 | 0.52 | (0.15, 1.81) | 0.305 |
| **Current clinical stage of HIV** |  |  |  |  |  |  |
| Stage I | 1.00 |  |  | 1.00 |  |  |
| Stage II | 0.87 | (0.59, 1.27) | 0.475 | 0.65 | (0.45, 0.94) | 0.021 |
| Stage III/IV | 0.48 | (0.36, 0.64) | <0.001 | 0.60 | (0.38, 0.95) | 0.030 |
| **Smoking** |  |  |  |  |  |  |
| Never smoked | 1.00 |  |  | 1.00 |  |  |
| Formerly smoked (>6 months) | 0.61 | (0.37, 0.99) | 0.044 | 0.83 | (0.56, 1.24) | 0.367 |
| Currently smokes (<6 months) | 0.53 | (0.26, 1.08) | 0.082 | 0.86 | (0.50, 1.48) | 0.595 |
| **Age** | 1.02 | (1.01, 1.03) | <0.001 | 1.00 | (0.99, 1.01) | 0.578 |
| **Duration on ART** | 1.03 | (1.01, 1.05) | <0.001 | 1.00 | (0.98, 1.02) | 0.949 |
| **Waist circumference** | 1.05 | (1.04, 1.06) | <0.001 | 0.99 | (0.96, 1.02) | 0.329 |
| **Hip circumference** | 1.03 | (1.02, 1.05) | <0.001 | 1.03 | (1.01, 1.05) | 0.005 |
| **Mid upper arm circumference** | 1.21 | (1.17, 1.24) | <0.001 | 1.10 | (1.05, 1.16) | <0.001 |
| **Neck circumference** | 1.11 | (1.07, 1.15) | <0.001 | 1.03 | (0.98, 1.07) | 0.209 |
| **Temperature** | 1.70 | (1.38, 2.10) | <0.001 | 1.15 | (0.94, 1.39) | 0.170 |
| **Systolic blood pressure** | 1.02 | (1.01, 1.02) | <0.001 | 1.00 | (0.99, 1.01) | 0.849 |
| **Diastolic blood pressure** | 1.02 | (1.02, 1.03) | <0.001 | 1.01 | (0.99, 1.03) | 0.215 |
| **Total Cholesterol (mmol/l)** | 1.28 | (1.15, 1.42) | <0.001 | 0.97 | (0.86, 1.09) | 0.588 |
| **LDL-Cholesterol (mmol/l)** | 1.30 | (1.15, 1.46) | <0.001 | 1.08 | (0.95, 1.24) | 0.238 |
| **Serum uric acid (mmol/l)** | 1.00 | (1.00, 1.00) | <0.001 | 1.00 | (1.00, 1.00) | 0.048 |
| **Waist hip ratio** | 3.69 | (1.24,11.00) | 0.019 | 24.69 | (0.97,628.17) | 0.052 |

**Table 2: Factors associated with obesity among people with HIV at an urban tertiary hospital in Uganda**

| **Characteristic** | **cPR** | **95% CI** | **P-value** | **aPR** | **95% CI** | **P-value** |
| --- | --- | --- | --- | --- | --- | --- |
| **TB history** |  |  |  |  |  |  |
| No | 1.00 |  |  | 1.00 |  |  |
| Yes | 0.31 | (0.18, 0.54) | <0.001 | 0.42 | (0.20, 0.88) | 0.021 |
| **Sex** |  |  |  |  |  |  |
| Male | 1.00 |  |  | 1.00 |  |  |
| Female | 3.46 | (1.94, 6.15) | <0.001 | 2.40 | (1.07, 5.38) | 0.034 |
| **Tribe** |  |  |  |  |  |  |
| Baganda | 1.00 |  |  | 1.00 |  |  |
| Banyakitara | 1.69 | (1.02, 2.78) | 0.040 | 1.14 | (0.72, 1.81) | 0.570 |
| Others | 1.08 | (0.55, 2.13) | 0.824 | 0.71 | (0.40, 1.28) | 0.257 |
| **Currently on Trimethoprim/Sulfamethoxazole prophylaxis** |  |  |  |  |  |  |
| No | 1.00 |  |  | 1.00 |  |  |
| Yes | 0.30 | (0.14, 0.68) | 0.004 | 0.55 | (0.24, 1.25) | 0.155 |
| **Current clinical stage of HIV** |  |  |  |  |  |  |
| Stage I | 1.00 |  |  | 1.00 |  |  |
| Stage II | 1.01 | (0.54, 1.90) | 0.973 | 0.45 | (0.23, 0.86) | 0.017 |
| Stage III/IV | 0.33 | (0.19, 0.58) | <0.001 | 1.29 | (0.61, 2.71) | 0.507 |
| **Age** | 1.04 | (1.02, 1.05) | <0.001 | 1.03 | (1.01, 1.05) | 0.011 |
| **Duration on ART** | 1.04 | (1.02, 1.07) | 0.002 | 0.98 | (0.94, 1.02) | 0.266 |
| **Waist circumference** | 1.08 | (1.06, 1.09) | <0.001 | 1.03 | (1.01, 1.05) | 0.010 |
| **Hip circumference** | 1.05 | (1.03, 1.06) | <0.001 | 1.03 | (1.02, 1.04) | <0.001 |
| **Mid upper arm circumference** | 1.34 | (1.28, 1.40) | <0.001 | 1.07 | (0.99, 1.17) | 0.083 |
| **Neck circumference** | 1.16 | (1.10, 1.23) | <0.001 | 1.10 | (0.99, 1.22) | 0.070 |
| **Temperature** | 2.22 | (1.52, 3.23) | <0.001 | 1.27 | (0.90, 1.79) | 0.179 |
| **Systolic blood pressure** | 1.02 | (1.01, 1.02) | 0.001 | 1.00 | (0.98, 1.01) | 0.610 |
| **Diastolic blood pressure** | 1.02 | (1.01, 1.04) | 0.001 | 1.01 | (0.98, 1.03) | 0.661 |
| **Total Cholesterol (mmol/l)** | 1.32 | (1.09, 1.61) | 0.005 | 0.86 | (0.70, 1.07) | 0.172 |
| **LDL-Cholesterol (mmol/l)** | 1.38 | (1.13, 1.67) | 0.001 | 1.08 | (0.85, 1.37) | 0.543 |
| **Serum uric acid (mmol/l)** | 1.00 | (1.00, 1.00) | 0.034 | 1.00 | (1.00, 1.00) | 0.297 |

**Table 3: Factors associated with central obesity among people with HIV at an urban tertiary hospital in Uganda**

| **Characteristic** | **cPR** | **95% CI** | **P-value** | **aPR** | **95% CI** | **P-value** |
| --- | --- | --- | --- | --- | --- | --- |
| **TB history** |  |  |  |  |  |  |
| No | 1.00 |  |  | 1.00 |  |  |
| Yes | 0.82 | (0.67, 1.01) | 0.058 | 1.00 | (0.83, 1.22) | 0.963 |
| **Sex** |  |  |  |  |  |  |
| Male | 1.00 |  |  | 1.00 |  |  |
| Female | 1.49 | (1.21, 1.84) | <0.001 | 1.78 | (1.28, 2.49) | 0.001 |
| **Marital Status** |  |  |  |  |  |  |
| Not Married | 1.00 |  |  | 1.00 |  |  |
| Married | 1.26 | (1.04, 1.54) | 0.020 | 1.29 | (1.05, 1.58) | 0.015 |
| **Education level** |  |  |  |  |  |  |
| Below Secondary | 1.00 |  |  | 1.00 |  |  |
| Secondary+ | 0.81 | (0.66, 0.99) | 0.036 | 0.95 | (0.78, 1.15) | 0.572 |
| **Dolutegravir-based ART** |  |  |  |  |  |  |
| No | 1.00 |  |  | 1.00 |  |  |
| Yes | 0.64 | (0.51, 0.81) | <0.001 | 0.98 | (0.76, 1.25) | 0.857 |
| **At least one past opportunistic infection (except LRTI)** |  |  |  |  |  |  |
| No | 1.00 |  |  | 1.00 |  |  |
| Yes | 1.30 | (1.03, 1.64) | 0.024 | 1.28 | (1.03, 1.59) | 0.027 |
| **Age** | 1.02 | (1.01, 1.03) | <0.001 | 1.02 | (1.01, 1.03) | 0.001 |
| **Duration on ART** | 1.03 | (1.02, 1.04) | <0.001 | 1.00 | (0.98, 1.02) | 0.808 |
| **Weight** | 1.02 | (1.02, 1.03) | <0.001 | 1.00 | (0.98, 1.01) | 0.765 |
| **Body Mass Index** | 1.07 | (1.06, 1.08) | <0.001 | 1.01 | (0.97, 1.05) | 0.533 |
| **Mid upper arm circumference** | 1.09 | (1.07, 1.11) | <0.001 | 1.04 | (0.99, 1.09) | 0.096 |
| **Neck circumference** | 1.05 | (1.02, 1.08) | <0.001 | 1.04 | (1.00, 1.08) | 0.049 |
| **Respiratory rate** | 1.05 | (1.01, 1.09) | 0.009 | 1.05 | (1.02, 1.09) | 0.004 |
| **Systolic blood pressure** | 1.01 | (1.01, 1.01) | <0.001 | 1.00 | (0.99, 1.01) | 0.586 |
| **Diastolic blood pressure** | 1.02 | (1.01, 1.02) | <0.001 | 1.01 | (0.99, 1.02) | 0.274 |
| **HBA1c** | 1.09 | (1.03, 1.15) | 0.002 | 1.06 | (1.00, 1.12) | 0.036 |
| **Total Cholesterol (mmol/l)** | 1.23 | (1.12, 1.35) | <0.001 | 1.01 | (0.92, 1.10) | 0.818 |
| **LDL-Cholesterol (mmol/l)** | 1.24 | (1.13, 1.36) | <0.001 | 1.12 | (1.01, 1.25) | 0.033 |
| **Triglycerides (mmol/l)** | 1.15 | (1.07, 1.23) | <0.001 | 1.09 | (0.99, 1.19) | 0.092 |
| **Serum uric acid (mmol/l)** | 1.00 | (1.00, 1.00) | 0.011 | 1.00 | (1.00, 1.00) | 0.255 |

**Table 4: Factors associated with hypertension among people with HIV at an urban tertiary hospital in Uganda**

| **Characteristic** | **cPR** | **95% CI** | **P-value** | **aPR** | **95% CI** | **P-value** |
| --- | --- | --- | --- | --- | --- | --- |
| **TB history** |  |  |  |  |  |  |
| No | 1.00 |  |  | 1.00 |  |  |
| Yes | 0.89 | (0.67, 1.19) | 0.427 | 0.82 | (0.60, 1.12) | 0.205 |
| **Dolutegravir** |  |  |  |  |  |  |
| No | 1.00 |  |  | 1.00 |  |  |
| Yes | 0.52 | (0.37, 0.72) | <0.001 | 0.96 | (0.62, 1.50) | 0.863 |
| **Family history of cardiovascular disease  in first degree relative** |  |  |  |  |  |  |
| No | 1.00 |  |  | 1.00 |  |  |
| Yes | 1.51 | (1.13, 2.02) | 0.005 | 1.29 | (0.96, 1.73) | 0.097 |
| **Age** | 1.04 | (1.03, 1.05) | <0.001 | 1.03 | (1.02, 1.05) | <0.001 |
| **Weight** | 1.02 | (1.02, 1.03) | <0.001 | 1.02 | (0.99, 1.05) | 0.117 |
| **Body Mass Index** | 1.05 | (1.03, 1.07) | <0.001 | 0.95 | (0.88, 1.01) | 0.115 |
| **Waist circumference** | 1.03 | (1.02, 1.04) | <0.001 | 0.99 | (0.95, 1.02) | 0.368 |
| **Hip circumference** | 1.02 | (1.01, 1.02) | <0.001 | 1.02 | (1.00, 1.04) | 0.017 |
| **Mid upper arm circumference** | 1.06 | (1.04, 1.09) | <0.001 | 1.01 | (0.94, 1.08) | 0.748 |
| **Neck circumference** | 1.10 | (1.06, 1.14) | <0.001 | 1.03 | (0.97, 1.09) | 0.385 |
| **Fasting blood glucose** | 1.12 | (1.03, 1.22) | 0.007 | 1.04 | (0.97, 1.13) | 0.282 |
| **HBA1c** | 1.16 | (1.08, 1.24) | <0.001 | 1.14 | (1.07, 1.21) | <0.001 |
| **Total Cholesterol (mmol/l)** | 1.30 | (1.14, 1.48) | <0.001 | 1.02 | (0.90, 1.16) | 0.722 |
| **HDL-Cholesterol (mmol/l)** | 1.53 | (1.27, 1.85) | <0.001 | 1.46 | (1.17, 1.82) | 0.001 |
| **Triglycerides (mmol/l)** | 1.15 | (1.05, 1.26) | 0.002 | 1.02 | (0.91, 1.14) | 0.741 |
| **Serum uric acid (mmol/l)** | 1.00 | (1.00, 1.00) | 0.025 | 1.00 | (1.00, 1.00) | 0.814 |
| **Waist hip ratio** | 5.68 | (2.14,15.08) | <0.001 | 11.42 | (0.88,149.09) | 0.063 |

**Table 5: Factors associated with diabetes mellitus among people with HIV at an urban tertiary hospital in Uganda**

| **Characteristic** | **cPR** | **95% CI** | **P-value** | **aPR** | **95% CI** | **P-value** |
| --- | --- | --- | --- | --- | --- | --- |
| **TB history** |  |  |  |  |  |  |
| No | 1.00 |  |  | 1.00 |  |  |
| Yes | 1.88 | (1.11, 3.17) | 0.018 | 2.34 | (1.11, 4.94) | 0.025 |
| **Dolutegravir** |  |  |  |  |  |  |
| No | 1.00 |  |  | 1.00 |  |  |
| Yes | 0.14 | (0.10, 0.22) | <0.001 | 0.19 | (0.11, 0.31) | <0.001 |
| **At least one current opportunistic infection** |  |  |  |  |  |  |
| No | 1.00 |  |  | 1.00 |  |  |
| Yes | 2.42 | (1.23, 4.74) | 0.010 | 2.70 | (1.35, 5.39) | 0.005 |
| **Current clinical stage of HIV** |  |  |  |  |  |  |
| Stage I | 1.00 |  |  | 1.00 |  |  |
| Stage II | 2.47 | (1.13, 5.41) | 0.024 | 2.00 | (0.95, 4.20) | 0.066 |
| Stage III/IV | 2.00 | (1.12, 3.57) | 0.020 | 0.88 | (0.42, 1.84) | 0.728 |
| **Family history of cardiovascular disease  in first degree relative** |  |  |  |  |  |  |
| No | 1.00 |  |  | 1.00 |  |  |
| Yes | 1.94 | (1.17, 3.24) | 0.011 | 1.66 | (0.99, 2.80) | 0.055 |
| **Age** | 1.04 | (1.02, 1.06) | <0.001 | 1.02 | (1.00, 1.04) | 0.071 |
| **Waist circumference** | 1.02 | (1.00, 1.03) | 0.016 | 1.00 | (0.99, 1.02) | 0.668 |
| **Systolic blood pressure** | 1.02 | (1.01, 1.03) | <0.001 | 1.01 | (0.98, 1.03) | 0.632 |
| **Diastolic blood pressure** | 1.02 | (1.01, 1.04) | 0.002 | 1.00 | (0.98, 1.03) | 0.736 |
| **HDL-Cholesterol (mmol/l)** | 2.01 | (1.53, 2.65) | <0.001 | 1.51 | (1.15, 2.00) | 0.003 |
| **Triglycerides (mmol/l)** | 1.24 | (1.08, 1.42) | 0.002 | 1.00 | (0.82, 1.22) | 0.997 |

**Table 6: Factors associated with elevated fasting blood glucose among people with HIV at an urban tertiary hospital in Uganda**

| **Characteristic** | **cPR** | **95% CI** | **P-value** | **aPR** | **95% CI** | **P-value** |
| --- | --- | --- | --- | --- | --- | --- |
| **TB history** |  |  |  |  |  |  |
| No | 1.00 |  |  | 1.00 |  |  |
| Yes | 1.82 | (1.39, 2.39) | <0.001 | 1.79 | (1.10, 2.92) | 0.020 |
| **Dolutegravir** |  |  |  |  |  |  |
| No | 1.00 |  |  | 1.00 |  |  |
| Yes | 0.65 | (0.46, 0.91) | 0.012 | 0.73 | (0.50, 1.07) | 0.108 |
| **At least one current opportunistic infection** |  |  |  |  |  |  |
| No | 1.00 |  |  | 1.00 |  |  |
| Yes | 1.92 | (1.42, 2.60) | <0.001 | 1.57 | (1.12, 2.19) | 0.009 |
| **Current clinical stage of HIV** |  |  |  |  |  |  |
| Stage I | 1.00 |  |  | 1.00 |  |  |
| Stage II | 0.70 | (0.36, 1.34) | 0.280 | 0.63 | (0.36, 1.12) | 0.113 |
| Stage III/IV | 1.57 | (1.20, 2.05) | 0.001 | 0.84 | (0.52, 1.34) | 0.458 |
| **Family history of cardiovascular disease  in first degree relative** |  |  |  |  |  |  |
| No | 1.00 |  |  | 1.00 |  |  |
| Yes | 1.38 | (1.07, 1.78) | 0.012 | 1.24 | (0.97, 1.59) | 0.082 |
| **Abnormalities on chest auscultation** |  |  |  |  |  |  |
| No | 1.00 |  |  | 1.00 |  |  |
| Yes | 1.54 | (1.10, 2.15) | 0.012 | 1.28 | (0.88, 1.86) | 0.190 |
| **Age** | 1.01 | (1.00, 1.02) | 0.040 | 1.00 | (0.99, 1.02) | 0.659 |
| **Systolic blood pressure** | 1.01 | (1.00, 1.01) | 0.002 | 1.00 | (0.99, 1.01) | 0.514 |
| **Diastolic blood pressure** | 1.01 | (1.00, 1.02) | 0.023 | 1.00 | (0.99, 1.02) | 0.889 |
| **Total Cholesterol (mmol/l)** | 1.14 | (1.00, 1.29) | 0.042 | 1.12 | (0.99, 1.26) | 0.074 |
| **HDL-Cholesterol (mmol/l)** | 1.55 | (1.30, 1.84) | <0.001 | 1.27 | (1.05, 1.54) | 0.013 |
| **Serum uric acid (mmol/l)** | 1.00 | (1.00, 1.00) | 0.027 | 1.00 | (1.00, 1.00) | 0.016 |
| **Waist hip ratio** | 3.02 | (1.16, 7.89) | 0.024 | 2.36 | (0.82, 6.83) | 0.113 |

**Table 7: Factors associated with elevated HbA1c among people with HIV at an urban tertiary hospital in Uganda**

| **Characteristic** | **cPR** | **95% CI** | **P-value** | **aPR** | **95% CI** | **P-value** |
| --- | --- | --- | --- | --- | --- | --- |
| **TB history** |  |  |  |  |  |  |
| No | 1.00 |  |  | 1.00 |  |  |
| Yes | 0.67 | (0.41, 1.07) | 0.093 | 0.77 | (0.47, 1.26) | 0.297 |
| **Hip circumference** | 1.02 | (1.00, 1.03) | 0.007 | 1.01 | (1.00, 1.03) | 0.029 |
| **SPO2 (%)** | 1.13 | (1.02, 1.25) | 0.022 | 1.11 | (1.01, 1.23) | 0.038 |
| **Systolic blood pressure** | 1.01 | (1.00, 1.02) | 0.009 | 1.01 | (1.00, 1.02) | 0.012 |

**Table 8: Factors associated with dyslipidemia among people with HIV at an urban tertiary hospital in Uganda**

| **Characteristic** | **cPR** | **95% CI** | **P-value** | **aPR** | **95% CI** | **P-value** |
| --- | --- | --- | --- | --- | --- | --- |
| **TB history** |  |  |  |  |  |  |
| No | 1.00 |  |  | 1.00 |  |  |
| Yes | 0.85 | (0.79, 0.91) | <0.001 | 0.92 | (0.83, 1.03) | 0.137 |
| **Sex** |  |  |  |  |  |  |
| Male | 1.00 |  |  | 1.00 |  |  |
| Female | 1.10 | (1.03, 1.18) | 0.010 | 1.12 | (1.02, 1.22) | 0.020 |
| **Current clinical stage of HIV** |  |  |  |  |  |  |
| Stage I | 1.00 |  |  | 1.00 |  |  |
| Stage II | 1.01 | (0.95, 1.08) | 0.641 | 0.99 | (0.93, 1.06) | 0.801 |
| Stage III/IV | 0.84 | (0.78, 0.91) | <0.001 | 0.93 | (0.83, 1.04) | 0.185 |
| **Alcohol use** |  |  |  |  |  |  |
| Never used alcohol | 1.00 |  |  | 1.00 |  |  |
| Formerly used alcohol (>6 months) | 1.01 | (0.95, 1.09) | 0.683 | 1.05 | (0.97, 1.13) | 0.204 |
| Currently uses alcohol (<6 months) | 0.89 | (0.81, 0.99) | 0.024 | 0.90 | (0.82, 0.98) | 0.023 |
| **Duration on ART** | 1.01 | (1.00, 1.01) | 0.002 | 1.00 | (1.00, 1.01) | 0.204 |
| **Weight** | 1.00 | (1.00, 1.01) | <0.001 | 1.01 | (1.00, 1.02) | 0.003 |
| **Body Mass Index** | 1.01 | (1.01, 1.02) | <0.001 | 0.99 | (0.97, 1.00) | 0.027 |
| **Waist circumference** | 1.00 | (1.00, 1.01) | <0.001 | 1.00 | (1.00, 1.00) | 0.822 |
| **Hip circumference** | 1.00 | (1.00, 1.01) | <0.001 | 1.00 | (0.99, 1.00) | 0.032 |
| **Mid upper arm circumference** | 1.01 | (1.01, 1.02) | 0.001 | 1.00 | (0.98, 1.02) | 0.820 |
| **Temperature** | 1.08 | (1.00, 1.15) | 0.038 | 1.06 | (0.99, 1.14) | 0.097 |

**Table 9: Ten-year cardiovascular risk among people with HIV with and without previously treated active TB in Uganda**

| **10-year CVD risk** | **Framingham score CVD risk categories*** | | |
| --- | --- | --- | --- |
|  | **No TB history n (%)** | **History of TB n (%)** | **p-value** |
| **Risk score (%**), median (IQR) | 1.2 (0.3 - 3.1) | 1.4 (0.4 – 4.3) | 0.180 |
| **Low risk** | 186 (93.5) | 169 (89.9) | 0.202 |
| **Moderate/High risk** | 13 (6.5) | 19 (10.1) |  |

CVD- cardiovascular disease, IQR – interquartile range, TB – tuberculosis. *Individuals were classified as having a low CVD risk if Framingham’s risk score (FRS) <10%; moderate to high CV risk if FRS is 10-20%; or high CV risk if FRS ≥20%.
